# Family Members’ Experiences Communicating with Coroners in Suspected Hereditary Sudden Cardiac Death Cases: A Mixed Methods Study

**DOI:** 10.1101/2025.10.10.25337778

**Authors:** Katherine S. Allan, Katherine L. Mason, Ariel Gershon, June Carroll, Katie N. Dainty, Joel A. Kirsh, Kris Cunningham, Paul Dorian, Arnon Adler, Julie Rutberg, Steve Lin, Sheldon Cheskes, Jodi Garner, Liz Siydock, Karolyn Yee, Krystina B. Lewis

## Abstract

**Background:** As SCD in the young can be caused by heritable cardiac conditions, up to 50% of family members may be at risk. Coroners are often responsible for communicating this risk, in addition to investigating the cause of death.

**Purpose:** To explore how family members of SCD victims experience the type, timing, and suitability of communication with coroners about the cause of death and their own risk for SCD.

**Methods:** We conducted an explanatory sequential mixed methods study. Eligible family members of SCD victims who died in 2021 from a potentially heritable cardiac condition and were investigated by the Office of the Chief Coroner of Ontario (Canada) were invited to complete a web survey and/or telephone interview. We used descriptive statistics to analyze the survey data and thematic analysis to analyze the interview data. Integration occurred at multiple levels.

**Results:** We sent survey invitations to 126 family members of 115 SCD victims; 50 family members completed the web-based survey between September 2022 and February 2024. We interviewed a subset of 17 family members. Most participants received initial communication by phone (n=30, 60%) or in-person (n=13, 26%). While many participants (n=39; 78%) received a timeline of when to expect death investigation results, 16 (32%) did not receive results within the expected timeframe. Family members characterized effective communication as clear, with coroners being perceived as kind, accessible, and approachable. When communication needs went unmet, families relied on external support networks to fill information gaps. Suggestions for improved communication included repeating information and using differing modes of communication to enhance understanding.

**Conclusions:** These findings emphasize the need for coroners to adopt empathetic approaches and to deliver information in a timely and clear manner. Addressing current gaps in communication may ensure the needs of grieving family members of SCD victims are met.

**What is Known:**

- The unexpected and sometimes unclear cause of sudden cardiac death (SCD), particularly in young persons, leaves families grieving, often with unanswered questions.
- In many jurisdictions, coroners investigate SCDs and have the most information regarding the circumstances and possible causes.
- Communication with coroners influences families’ experiences with respect to learning about the cause of death and their own personal risk for a heritable cardiac condition.

**What this Study Adds:**

- Family members of SCD victims seek clarity, accessibility, approachability, and kindness in their interactions with coroners.
- When family members’ communication needs went unmet, many independently sought resources and support networks outside the coroner system.
- Family members recommend improving coroner communication by providing clear timelines for death investigation results, repeating key information using several different modes of communication, and directing the family to counseling resources.

## Background

Sudden cardiac death (SCD) is a prevalent cause of mortality in North America, with an incidence of approximately 50-100 per 100,000.^1^ Due to its unexpected nature, it is a devastating event for both families and the community.^2,3^ SCD is defined as a natural event with cardiorespiratory collapse, which occurs suddenly, unexpectedly and is presumed to be due to a cardiac cause.^4^ There are diverse underlying conditions that can lead to SCD, including both acquired and heritable causes.

Inherited arrhythmia syndromes are much more common in younger (age < 50) people. In circumstances where the death is sudden and of an unknown cause, international guidelines recommend an autopsy to help determine the underlying reason for the SCD, and whether it may be from an inherited cardiac condition.^5^ Determining if the cause of death is hereditary is critical, as up to 50% of first-degree family members may be at risk.^6^ Additionally, a diagnosis of cardiac disease in the decedent can inform family members of their potential future risk of heart disease.^7^

According to Ontario (Canada) provincial legislation, coroners are responsible for investigating and determining the cause of sudden, unexpected, and/or unexplained deaths and initiating and maintaining contact with the family throughout the investigation.^8^ The communication and conversations that occur throughout the death investigation are complex. In the context of a sudden and devastating event for surviving family members, coroners must present and describe complex medical information, while family members experience heightened emotions, and many of whom experience symptoms of post-traumatic stress, depression, anxiety, and prolonged grief.^9^ Moreover, as coroners typically communicate with one primary contact during the death investigation, the communication between them has implications for what is shared and how it is shared with other family members. How coroners communicate with families throughout this death investigation can influence next-of-kin’s understanding of the cause of death, of their own risk for a heritable cardiac condition, and whether appropriate follow-up, diagnoses, and treatment are perceived as needed, sought, and received for the people affected. We have previously explored the experiences of family members throughout the death investigation process; yet the ways in which communication occurs between coroners and family members is still not well understood.^2,10^

This paper explores how family members of SCD victims perceive and experience communication with coroners throughout the death investigation process. Three research questions guided this study:

1. How do family members rate the type, timing, and appropriateness of communication strategies used by coroners during the death investigation process (quantitative)?
2. What are the experiences of family members communicating with coroners regarding the cause of death and their potential risk for SCD (qualitative)?
3. How do family members’ experiences and perceptions of coroner communication align with their needs and preferences for such interactions (mixed methods)?

## Methods

### Study Design

We conducted a sequential explanatory mixed methods study in two phases to comprehensively explore family members’ perceptions and experiences of communication with coroners following SCD.^11^ In this context, we considered the communication process as measurable (in terms of timing, type, and perceived appropriateness) and deeply personal, involving emotional, social, and subjective dimensions that require contextual understanding. The integration of both data allowed for a richer, more comprehensive understanding of the issue than either method could provide alone. First, we administered a web-based quantitative survey to family members of SCD victims followed by semi-structured interviews with a subset of web-survey participants. The University of Ottawa Research Ethics Board approved this study (H-09-21-7135). We report this study using the CHERRIES,^12^ SRQR^13^ and GRAMMS^14^ reporting guidelines.

### Research Characteristics and Reflexivity

Our research team included members of the Office of the Chief Coroner of Ontario (K.C., J.A.K., L.S.) and the Genetic Condition Coordinator (K.S.A), who were seeking evidence-based strategies to improve communication between coroners and the families of SCD victims during the death investigation process. In the absence of available evidence, the team was interested in hearing directly from family members who had navigated the Ontario death investigation system in the past 4 years, to design and develop strategies rooted in their experiences. A family member partner (J.G.) who had lost a family member to SCD, has been driven to improve the timeliness of the SCD investigation for others. Our research team included early, mid and senior career investigators with mixed methods experience (K.B.L., K.S.A., K.N.D.), trainees (K.L.M, A.G., K.Y.), a genetic counsellor (J.R.) and clinician-researchers in cardiology (A.A., P.D.), family and emergency medicine (J.C., S.C., S.L.), all with the knowledge and desire to improve coroner communication for families who have recently lost a loved one to SCD. Working in collaboration with these knowledge users to integrate research findings into practice, is known as an integrated knowledge translation approach.^15^

### Setting

This study was conducted in Ontario, Canada, which has the largest death investigation system in North America.^16^ Legislation mandates that coroners investigate all sudden and unexpected deaths to determine the cause and manner of death. Death investigations generally involve collecting full medical and social histories, ordering a post-mortem examination, including toxicology, and requesting post-mortem genetic testing as required. Coroners are also responsible for supporting families throughout the death investigation and informing them of the final investigation results and implications. Coroners typically only communicate with one member of the victim’s family, who is designated as the primary point of contact for communicating to the rest of the family and other interested parties.

### Eligibility Criteria

We used an existing standardized screening process to identify all SCDs caused by heritable cardiac conditions (i.e. inherited arrhythmia or heritable cardiomyopathy) by reviewing death investigation files (e.g., coroner’s investigation statement, post-mortem report, toxicology, post-mortem genetic testing results).^2,17^ Deaths were considered eligible if they were investigated by a coroner, were caused by a potentially heritable cardiac condition as per the Canadian Cardiovascular Society Guidelines,^18^ occurred in Ontario in 2021 and were between the ages of 2-70. We chose age 2 as the lower age cut-off to reduce overlap with sudden infant death syndrome; and 70 years as the upper age cut-off, as cardiomyopathies can affect individuals at any age.^19^ Participating family members were required to be 18 and over, and able to read, understand, and speak either French or English.

### Recruitment

For each eligible SCD case, we informed the primary next-of-kin about the study by email and invited them to participate using a secure link to the anonymous web survey. In the cases where victims had more than one next-of-kin identified, we invited all to participate. Despite only contacting the primary next-of-kin, we had no control over how many family members per decedent responded, as next-of-kin could share the study information and secure web-link with others. We sent two follow-up emails, each 2 weeks apart, to all eligible next-of-kin. We invited participants who completed the web survey to participate in a telephone interview to ensure continuity in our sampling approach. If interested, they were contacted by the research team to arrange an interview timeslot. We sent one follow-up email one week later to non-respondents for the telephone interview.

## Data collection

### Quantitative data collection

We collected quantitative data through a web survey, using Hosted in Canada Surveys, between September 2022 and February 2024. Completion of the web-based survey implied consent. We developed the web survey based on previous research, known gaps in healthcare communication, and our team’s expertise in the SCD field.^2,20^ Prior to its launch, family members of SCD victims not meeting study eligibility criteria piloted the web survey and we made minor edits for comprehensibility and flow. The web survey included 84 questions, distributed one page at a time. Questions were organized in 9 sections, comprised of mostly closed-ended questions, with some open-ended questions to offer further explanation for responses. See Supplemental Appendix for details. We provided web survey participants who chose to provide their email address with a $20 electronic gift card. We carefully reviewed email addresses for duplicates.

### Qualitative data collection

We collected qualitative data via telephone interviews with a subset of survey respondents. We developed a semi-structured interview guide, informed by the web survey results and prior research,^2,17,20^ aiming to further explain survey responses and enhance the relevance, focus, and depth of the qualitative inquiry. K.B.L. and K.L.M conducted telephone interviews between September 2023 and February 2024. Verbal consent was obtained at the beginning of telephone interviews. Interviews were audio recorded, professionally transcribed and de-identified at the time of transcription. Interview participants were offered an additional $20 electronic gift card for completing the interview.

## Data Analysis

### Quantitative data analysis

We used descriptive statistics to analyze all quantitative data. We report binary data as counts and percentages, while continuous data are reported as means and standard deviations. Participants were required to complete at least two survey sections (of 9) to be included in the analysis. We report the number of incomplete responses for each question.

### Qualitative data analysis

Our team of two PhD-prepared researchers (K.S.A. and K.B.L.) and three trainees (K.L.M, K.Y., and A.G.) engaged in a team-based inductive thematic analysis^21,22^ process to identify and organize meaningful segments of data that arose from the interview transcripts. Two team members independently analyzed each transcript. At bi-weekly team meetings, the analysis team discussed the coding, the development and evolution of the codebook, and the evolution of themes. The Microsoft Excel based codebook was continuously revised to reflect evolving codes, patterns, and themes. With a final version of the codebook agreed upon by the analytical team, all interview transcripts were re-analyzed to ensure relevant data was captured.

### Data integration

We integrated qualitative and quantitative data at several levels: study design, methods, data collection and interpretation. At the study design level, we used an explanatory sequential design, where the results from the web-based quantitative survey informed, expanded, and explained qualitative data collection. At the methods level, we recruited a subset of participants who completed the web-based survey to participate in interviews which ensured continuity in our sampling approach. We used the results from the web survey to adjust our interview guide to more deeply explore the patterns of family members’ communication with coroners revealed during the survey, ensuring that our interview questions were responsive to real-world experiences. At the interpretation and reporting level, we integrated data from both the web-survey and telephone interview using joint displays and weaving techniques in the presentation of our results.^23^

## Results

### Participant characteristics

Of the 1488 death records presumed to be caused by heritable cardiac conditions in the database in 2021, 115 deaths met our eligibility criteria (Figure 1). We contacted 126 family members of these 115 decedents. Of these, 50 family members agreed to participate and completed at least 2 sections of the web survey (40% response rate). The mean age of survey participants was 48 +/- 14 years old, and 35 (70%) self-identified as women, while 36 (74%) were female sex at birth. About half of respondents were either a parent or sibling to the decedent (Table 1).

**Figure 1:**
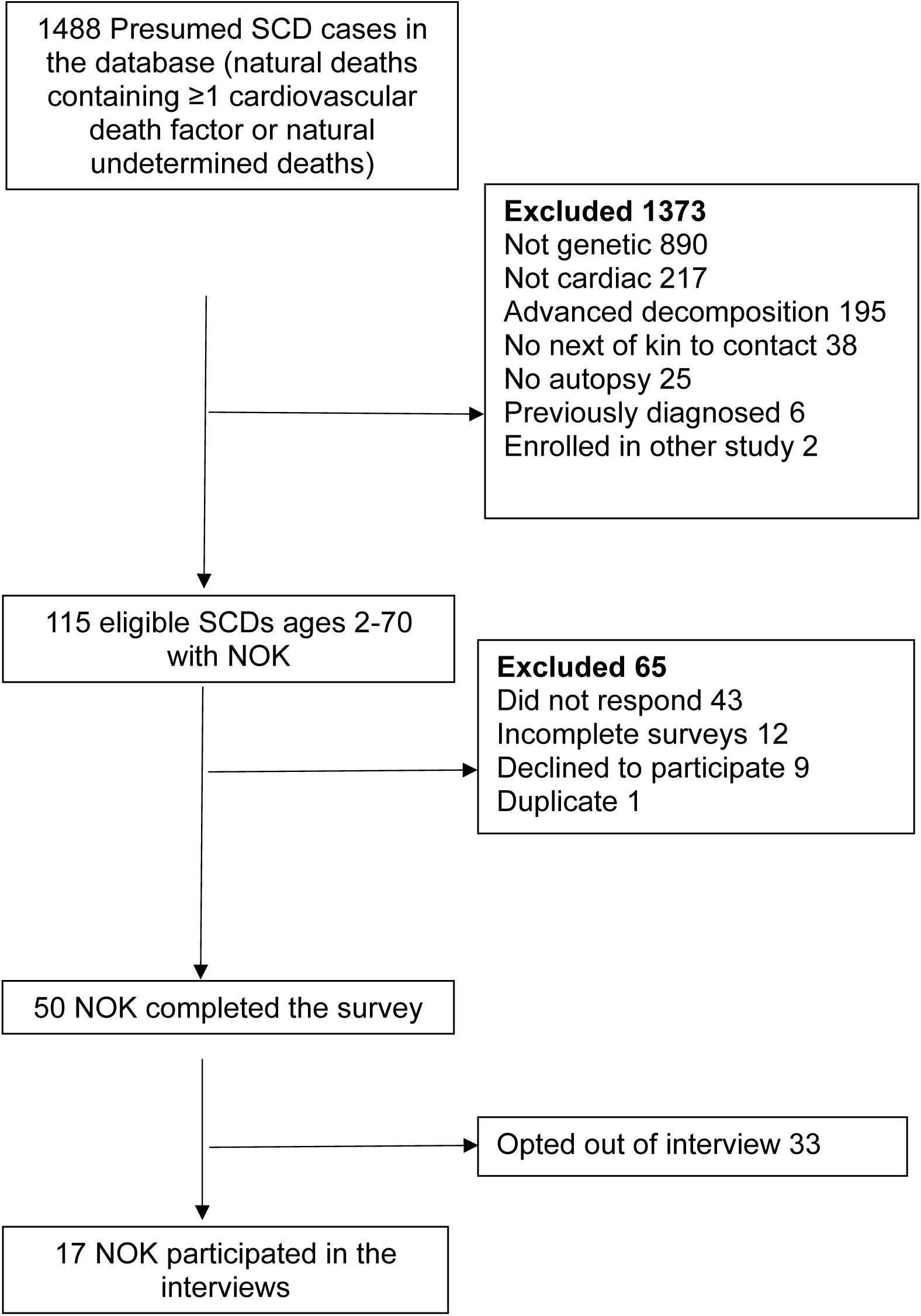
Participant flow diagram.

**Table 1.**
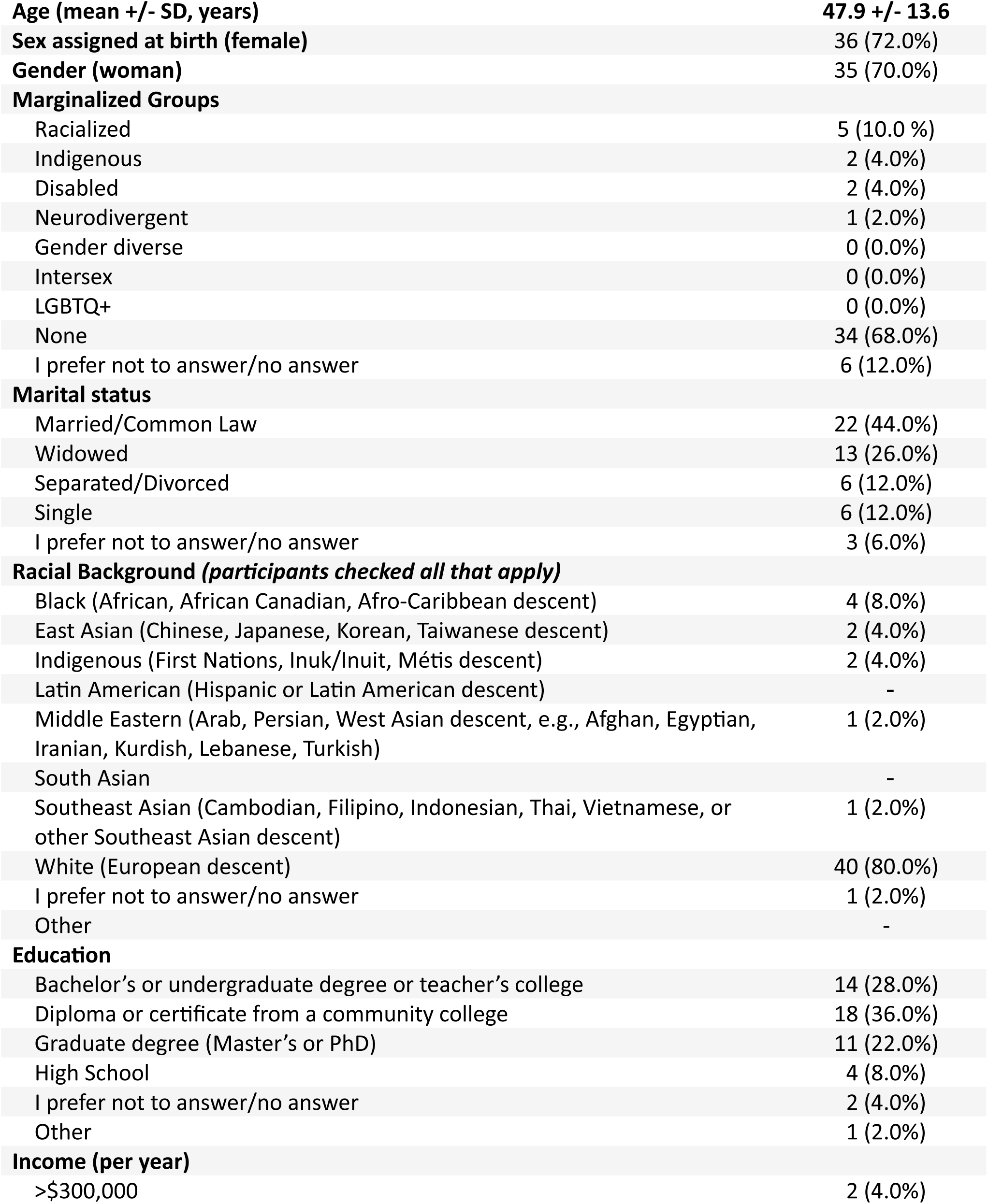
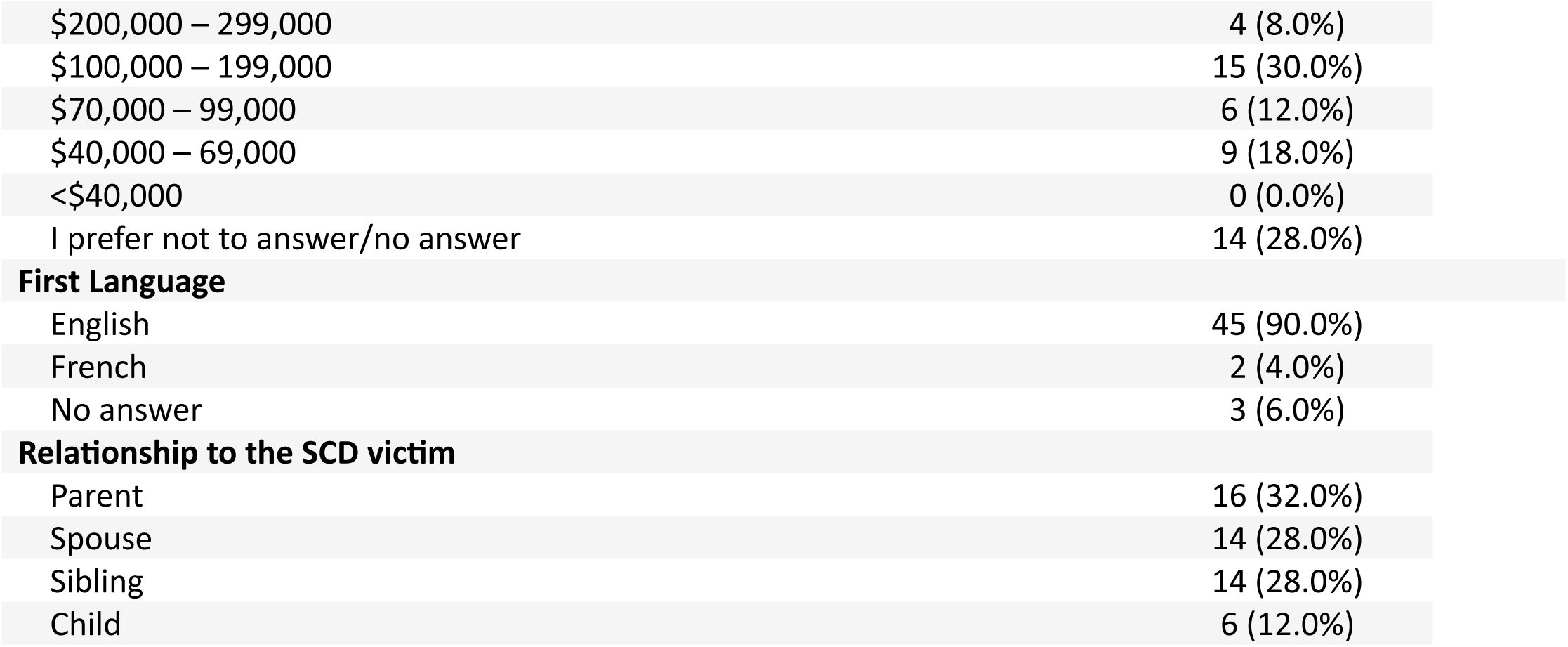
Participant characteristics (n=50)

Survey participants provided demographic information about their deceased family member. The mean age of SCD victims was 38 +/- 12 years old, with 11 (22%) women. Respondents attributed the death to a variety of causes, with 12 (24%) reporting that they did not know the cause of death and nineteen of 50 (38%) reported the cause of death as a cardiomyopathy (Table 2).

**Table 2.** Characteristics of SCD victims (n=50) as reported by their family members.

|  |  |
| --- | --- |
| **Age (mean +/- SD, years)** | 38.7 +/- 11.5 |
| **Sex** |  |
| Male | 39 (78.0%) |
| Female | 11 (22.0%) |
| Intersex | 0 (0.0%) |
| Prefer not to answer/no answer | 0 (0.0%) |
| **Gender** |  |
| Man | 39 (78.0%) |
| Woman | 11 (22.0%) |
| Prefer not to answer/no answer | 0 (0.0%) |
| Other | 0 (0.0%) |
| **Do you know the cause of death?** |  |
| Yes, I know | 38 (76.0%) |
| No, I don't know | 11 (22.0%) |
| Unsure | 1 (2.0%) |
| **Cause of death** |  |
| Aortopathy |  |
| Aortic rupture, dilation, or dissection | 8 (16.0%) |
| Cardiomyopathy |  |
| Cardiomyopathy (not otherwise specified) | 1 (2.0%) |
| Hypertrophic Cardiomyopathy (HCM) | 2 (4.0%) |
| ARVC | 3 (6.0%) |
| Dilated Cardiomyopathy | 13 (26.0%) |
| Peripartum Cardiomyopathy | 2 (4.0%) |
| Arrhythmia Syndrome (LQTS, Brugada Syndrome, CPVT, Short QT) | 1 (2.0%) |
| Coronary Artery Disease | 3 (6.0%) |
| I can't remember | 2 (4.0%) |
| Other* | 6 (12.0%) |
| No answer | 11 (22.0%) |

Abbreviations: ARVC: Arrhythmogenic Right Ventricular Cardiomyopathy; CPVT: Catecholaminergic polymorphic ventricular tachycardia; LQTS: Long QT Syndrome

Of the 50 web survey participants, 17 (34%) agreed to participate in a telephone interview. In one SCD case, two family members participated in the telephone interview. The mean age of interview participants was 48 +/- 12 years, and the majority self-identified as women (14; 78%) (Table 1).

### Quantitative Web Survey Results

Web survey findings revealed that 17 (34%) family members were contacted by the coroner the day of their family member’s death, 7 (14%) were contacted the next day, and 14 (28%) were contacted between 24 hours and 1 week after the death (Table 3). Over half (n=30; 60%) were initially contacted by telephone, while 13 (26%) were approached in person. Most family members were extremely (n=18; 36%) or very satisfied (n=17; 34%) regarding the mode of contact that was used.

**Table 3.** Family members’ perceptions of coroner communication (n=50)

|  |  |  |
| --- | --- | --- |
| During the period following your family member's death, did you have any contact with the coroner? |  |  |
| Yes | 48 (96.0%) | *"I had fairly early communication with the Coroner and then they contacted me shortly after with further information" (P-13)* |
| No | 2 (4.0%) |  |
| How long after the death of your family member did the coroner contact you? |  |  |
| The same day | 17 (34.0%) | *"I'm not sure if it was the same day or the next day that I first talked to him" (P-15)* |
| The next day | 7 (14.0%) |  |
| Between 24 hrs and 1 week | 14 (28.0%) |  |
| Between 1 week and 1 month | 3 (6.0%) |  |
| More than one month | 5 (10.0%) |  |
| I don't recall | 4 (8.0%) |  |
| How did the coroner initially contact you? |  |  |
| Email | 5 (10.0%) | *"I was at the apartment, and we waited for the coroner to arrive. We had to wait a little while. I understand, that wasn't an issue. He introduced himself. He took us aside, he explained to us, he told us who he was." (P-17)* |
| In-person | 13 (26.0%) |  |
| Telephone/Voicemail | 30 (60.0%) |  |
| Written letter sent by post | 1 (2.0%) |  |
| I don't recall | 1 (2.0%) |  |
| Were you satisfied with the method of contact that the coroner used? |  |  |
| Extremely satisfied | 18 (36.0%) | *"I think it was important for the first conversation [to be] on the phone, and you know, she took all the time around the situation. I thought that was great." (P-06)* |
| Very satisfied | 17 (34.0%) |  |
| Neutral | 12 (24.0%) |  |
| Not very satisfied | 1 (2.0%) |  |
| Not at all satisfied | 2 (4.0%) |  |
| After the initial contact, did you have any further contact with the coroner? |  |  |
| Yes | 44 (88.0%) | *"Whenever there was any information to be shared, he contacted me." (P-05)* |
| No | 3 (6.0%) |  |
| I don't recall | 3 (6.0%) |  |
| Frequency of communication |  |  |
| 1 to 5 times | 37 (74.0%) | *"They were pretty minimal. There was just the initial time at the hospital and then* |
| 6 to 10 times | 7 (14.0%) | there's a couple, she did a follow up call maybe a week later." (P-04) |
| Not applicable | 6 (12.0%) |  |
| Did the coroner return your calls and/or respond to your emails/letters in a timely manner? |  |  |
| Yes | 38 (76.0%) | "She was always like, whenever you're available, whenever you want to speak with me. It's kind of an open door." (P-06) |
| No | 5 (10.0%) |  |
| I don't recall | 7 (14.0%) |  |
| When you had questions for the coroner, did you receive answers in a way that you easily understood the information? |  |  |
| Yes | 40 (80.0%) | "She broke it down like explaining the heart and what had happened and what didn't happen. Yeah, so I felt like I understood that." (P-03) |
| No | 6 (12.0%) |  |
| I don't recall | 4 (8.0%) |  |
| Did the coroner provide you with a timeline for when you might receive results? |  |  |
| Yes | 39 (78.0%) | "I do remember that taking like quite a few months to receive, which we had been told that that was going to be the case." (P-01) |
| No | 5 (10.0%) |  |
| I don't recall | 4 (8.0%) |  |
| No answer | 2 (4.0%) |  |
| Do you recall if you received the results from the death investigation process within the timeframe that was described to you? |  |  |
| Yes | 25 (50.0%) | "I think what was helpful was everything that they said they were going to do, they did do, and they did provide me with information." (P-17) |
| No | 16 (32.0%) |  |
| I don't recall | 8 (16.0%) |  |
| No answer | 1 (2.0%) |  |

Forty-seven (94%) participants reported receiving information about the cause of death from the coroner. Most participants felt that the information they received was helpful (n=33, 66%), while others reported that the information was not (n=6, 12%). Five participants (10%) reported that the amount of information received was not enough. Most family members (39; 78%) recalled receiving a timeline for the death investigation results, yet 16 (32%) did not recall receiving the results within the anticipated timeframe. When family members were provided with results within the estimated timeline, they described being extremely satisfied (5; 10%), very satisfied (12; 24%), or neutral (11; 22%).

Thirteen (26%) family members never received information about the cause of death from the coroner. For those who did receive information about the cause of death, most reported feeling extremely satisfied (n=5, 10%), very satisfied (n=19, 38%), or neutral (n=12, 24%) about the explanation the received. One participant (2%) reported feeling not at all satisfied with the explanation they received.

### Qualitative Telephone Interview Results

Through our inductive analysis, we revealed two main themes and several subthemes. Theme 1 focuses on the *qualities of good coroner communication* from the perspective of families. Theme 1 included two subthemes, which revealed that good communication is characterized by its clarity (subtheme 1) and by the coroner’s demeanor, specifically about being accessible, approachable and kind (subtheme 2). Theme 2 describes *system-level constraints to good communication between coroners and families*. These system-level constraints were primarily about the timing of communication and ability to meet expectations (subtheme 1); filling system gaps by seeking outside resources (subtheme 2), and proposed solutions to enhance communication between coroners and families (subtheme 3).

### Theme 1. Qualities of good coroner communication

#### Subtheme 1. Being clear

All family members who were interviewed valued clear communication from the coroner, which influenced their level of understanding of their loved one’s cause of death and their own risk of heritable cardiac conditions. Family members described having difficulty understanding and absorbing information, processing their emotions throughout their grief, and trying to piece together the events leading up to the SCD event. When communication was clear, family members described information exchanges that were easy for them to understand. In these instances, coroners would use plain and simple language. One family member described that the coroner “*was good at explaining what things meant. Because the reports were very complicated. So, [the coroner] would break it down into terms that [the family member] could relate to”* (Participant 04).

Some interview participants described that the explanation of the cause of death was insufficiently detailed, leading to frustration and confusion. In other cases, coroners provided explanations using complex language or provided a complicated explanation for the cause of death, which some family members struggled to comprehend or absorb. One family member described that they “*had no idea what it meant to have dilated cardiomyopathy”* and *“it was definitely not in layman’s terms whatsoever”* (Participant 02).

#### Subtheme 2. Being accessible, approachable and kind - The coroner’s demeanor

Most participants valued the coroner’s demeanor, describing how their overall manner shaped family members’ experiences communicating with them. Coroners with overall positive demeanours were considered approachable, and families were more comfortable reaching out to them. In many cases, family members stated that it was helpful when the coroner was available and willing to answer their questions. One family member spoke of this, saying “*[the coroner] was available, he was super friendly, he was down to earth, he was open to answering any of my questions, even when I knew they were ridiculous … If I ever need anything, any questions, I can call him anytime”* (Participant 8).

When family members reported a positive impression of the coroner, they described them as respectful, supportive, understanding, polite, and prioritizing their case. One family member described that they *“appreciated that [the coroner] made the time, and [they] felt prioritized when [the coroner] reached out, even though it took [the family member] a year, [they] still felt prioritized”* (Participant 06). Family members valued moments where the coroner’s demeanour was kind, compassionate, and warm.

In other cases, family members had an overall negative impression of the coroner. In these instances, family members described them as abrupt, lacking empathy and understanding, or behaving coldly. One family member described *“I don’t remember it being a very positive experience because he didn’t give off the impression of providing any sort of empathy or understanding on what it was that we were going through”* (Participant 01).

### Theme 2: System-level constraints to good communication

#### Subtheme 1. Timing and ability to meet expectations

The type, timing, and frequency of communication between coroners and family members of SCD victims varied and was often dependent on the circumstances of the case. A few family members described the value of in-person communication because they appreciated the *“human element, to have someone come to my house to have that face-to-face conversation”* because *“it’s a really difficult thing to speak to over the phone”* (Participant 01). Despite this preference, the survey results indicated that in-person communication was a less common modality than phone calls and emails, and most expressed no issue with the method of communication chosen.

Timing of the first communication from the coroner was typically between the *“initial time at the hospital”* (Participant 04) and *“two or three days after”* (Participant 01) the death, depending on the circumstances of the case. Family members spoke of long wait periods before receiving an official autopsy report, sometimes waiting *“almost six months”* (Participant 06) or receiving final reports *“almost a year later”* (Participant 17).

Participants also referred to the coroner’s ability to set and meet expectations for the death investigation. Family members appreciated when the coroner described next steps and how the death investigation would proceed. Family members valued the moments when the coroner met these expectations, they had set or promptly communicated any changes in the investigation process with the family. Conversely, some coroners did not set expectations or did not communicate with family members along the timeline they had promised, which negatively impacted the family’s experience. One family member said that *“it would have been nice if someone told [them] that this stuff doesn’t happen right away and [they] could be waiting months, and to prepare for that”* (Participant 7).

#### Subtheme 2. Filling coroner system gaps by seeking outside resources

Communication between coroners and family members of SCD victims did not happen in isolation. Family members reported that their personal circumstances and networks provided access to additional resources, which enhanced their communication with the coroner. For example, some family members described how their own level and field of education, as well as their professional background, influenced their ability to understand the content. One’s personal support circle and the value of having *“a second person with [them]”* (Participant 08) while communicating with the coroner was also deemed advantageous.

Some participants spoke of the additional usefulness of speaking with *“friends and relatives that are doctors”* (Participant 15) to help them decipher the coroner’s information. Others were not as fortunate, describing that they *“didn’t feel like [they] had anybody”* (Participant 02) that they could reach out to beyond the coroner, to answer their questions or access additional information. In most cases, the family physician of the participant was included in the death investigation process, typically by providing referrals to cardiologists for cardiac screening. In some cases, this led family physicians to offer to review the autopsy report with family members.

#### Subtheme 3. Proposed solutions to enhance communication between coroners and families

To improve the clarity of communication, participants suggested repeating the same information using different modes of communication to enhance understanding. This could include using both verbal and written communication at different times during the death investigation process. Additionally, a family member suggested that coroners allow 24 hours between the SCD and their initial communication to allow families extra time to process the tragic circumstances. A few participants recommended that coroners provide family members with a warning before sharing investigation results, updates, or autopsy reports, to help prepare them to receive such difficult information. One participant described that *“there’s really no heads-up as to when [the autopsy report is] coming. So, you could finally be having an okay day and get outside and then you’d check your mail, and the next thing you know you’re holding a report in your hand that tells you how much your brother’s heart weighed”* (Participant 02).

Family members described a need for increased frequency of communication with coroners. One participant described *“wishing that [they] had gotten more updates along the way”* (Participant 03). Some participants described the helpfulness of shifting certain tasks related to identifying and submitting the necessary paperwork to the coroner. Another participant suggested including an online portal where family members could check the status of the coroner investigation and autopsy report.

Many participants mentioned accessing external resources independently to seek further information. One family member recommended that coroners provide the family with a list of relevant resources, such as therapy and grief counselling following the SCD *“because it took a lot of energy just to fill out the intake forms”* (Participant 03). Another participant described the value of creating a tool for family members *“to easily streamline [referrals]. So, [they’re] not having to go hunt everything down yourself”* (Participant 04).

## Data Integration

There was convergence between the quantitative and qualitative data on what constituted the qualities of good coroner communication and how the coroner’s demeanor influenced families’ experiences. While the survey measured how many families received information clearly, how often and by what means, the qualitative interviews expanded on these topics to explore *how* the coroners were clear in their communication (and when they weren’t) and *how* their overall demeanor shaped the experience, positively or negatively (complementarity). When communication needs were unmet or were negatively perceived, the qualitative interviews expanded on how families coped, which included seeking outside resources and providing suggestions on how to improve. These integrated results are presented in a joint display and meta-inferences table, with the survey results juxtaposed with representative quotes (Table 3 and Supplemental Table 1).

## Discussion

This mixed methods study showed that most family members were satisfied with the type and timing of communication used by coroners during the SCD death investigation of their relative. Almost half of family members were first contacted by coroners within 24 hours of their loved one’s death and the most frequent mode of communication used was by telephone. Family members characterized effective communication as clear, with coroners being perceived as kind, accessible, and approachable. When their communication needs were unmet or were unsatisfactory, family members sought outside resources to fill these gaps. Many family members proposed solutions to improve the communication experience.

In our study, the survey results revealed that the most frequently used mode of communication was by telephone. As the majority of the SCDs included in this study occurred during 2021, it is likely the COVID-19 pandemic influenced a change from in person to virtual communication to abide by provincial guidelines.^25^ As heard from our knowledge users on the research team, this shift to phone/virtual communication has remained since, despite permissions to return to in-person interactions. In another study conducted by our team, coroners reported that SCD cases are more difficult to investigate, require more frequent communication, and as such do benefit from using different modes of communication modalities - yet admitted to using the telephone most often.^25^ Coroners’ choice of modality depended on their personal preferences, the circumstances of the investigation, and the preferences of the family.^25^

Our integrated results highlighted that an important factor for influencing how communication with coroners was perceived by family members, was the early establishment of an estimated timeline for the death investigation. When family members were provided with results within this timeline, they expressed satisfaction; those that did not receive results within this timeline expressed negative experiences. Numerous family members spoke of long wait times (6-12 months) before receiving an official autopsy report. This is concerning, as families affected by SCD greatly desire a clear medical explanation for the cause of death,^26–28^ thus long waits for information can lead to them becoming increasingly anxious and distressed and overly prolong the grieving process.^2,29^ Open lines of communication between family members and coroners may mitigate the negative implications of long wait times for autopsy reports by appropriately setting family member’s expectations. Coroners should attempt to provide clear, straightforward explanations of the cause of death as soon as the results are available and recognize that this conversation may need to be revisited several times.

Qualitative interviews with families provided greater depth about the qualities of good communication between families and coroners, that goes beyond the spoken word and revealed the importance of the coroner’s demeanor. This is consistent with other studies on neonatal and pediatric deaths, which have shown that parents appreciate healthcare professionals who show emotion, while those who lack emotion are perceived as being cold and uncaring.^30,31^ There are also many other factors that can influence a family members’ ability to communicate and comprehend the complex information shared by coroners during a death investigation. Family members of SCD victims described how their educational and professional background influenced their ability to comprehend information communicated to them by coroners. Highly educated family members, or those with a relevant health or scientific education or professional background, may understand this information more easily than family members who do not have these backgrounds. Individuals with higher health literacy, a key social determinant of health,^33^ are also more likely to show interest and to participate in research.^32^ Individuals with higher health literacy may better understand their own risk of heritable cardiac condition and in turn, be better able to mitigate that risk. Conversely, barriers to engaging with the healthcare system could cause some to suffer worse health outcomes due to missed opportunities to screen for and treat preventable conditions. Thus, coroners may wish to ask family members about their educational status, familiarity, and comfort with the healthcare/medical field, and tailor their approach and communication accordingly.

## Limitations

Our study is not without limitations. First, our study occurred in Ontario’s coronial system, where all coroners are medical doctors by training. The communication dynamics may be different in other coroner systems, where coroners have nursing or legal backgrounds, or no medical training at all. Second, our study sample included a higher proportion of participants who self-identified as women for both phases. This distribution may reflect that death from cardiac causes is more common in men or that perhaps women are more inclined to share their experiences. Third, over two-thirds of our survey sample received post-secondary education. The findings of this study may not be applicable to populations with lower health literacy. Finally, there is an important proportion of “no answer” responses to some survey questions. Despite piloting our survey with family members of SCD victims, it is possible that some participants found the survey long, uncomfortable, or difficult to answer. This is an important reminder that care must be taken when conducting research with this vulnerable group.

## Conclusion

Our findings demonstrated that most family members were satisfied with the initial mode of communication, as well as the timing of communication used throughout the death investigation. Family members valued clarity of communication as well as the coroner’s demeanor. Our findings emphasize the need for coroners to adopt empathetic approaches and deliver information clearly to family of SCD victims. Family members provided numerous suggestions on how to improve communication with coroners, such as repeating the same information using different modes of communication, and providing a warning before sending investigation results. Addressing current gaps in communication may ensure the needs of grieving family members of SCD victims are met.

## Data Availability

The data that support the findings of this study are available on request from the corresponding author, [KSA]. The data are not publicly available due to privacy and ethics restrictions.

## Nonstandard Abbreviations and Acronyms

SCD: Sudden Cardiac Death

## Acknowledgements

The authors wish to thank the families of sudden cardiac death victims for their time and for their willingness to share their experiences with us. We would like to acknowledge Andrew Stephen and Dr. Dirk Huyer, from the Office of the Chief Coroner of Ontario for their support and involvement in this study.

